# Prediction of vasospasms as complication in ischemic stroke patients receiving anterior circulation endovascular stroke treatment

**DOI:** 10.1101/2022.12.18.22283313

**Authors:** Jessica Jesser, Sinclair Awounvo, Johannes A. Vey, Dominik Vollherbst, Tim Hilgenfeld, Min Chen, Silvia Schönenberger, Martin Bendszus, Markus A. Möhlenbruch, Charlotte S. Weyland

## Abstract

**Background and Purpose:** Vasospasms are a common and dreaded complication of endovascular stroke treatment (EST). There is a lack of understanding of risk factors for periprocedural vasospasms. Here, we aimed to identify prognostic factors for vasospasms in patients with acute ischemic stroke and EST.

**Methods:** Retrospective single-center analysis of patients receiving EST for anterior circulation vessel occlusion between January 2015 and December 2021. Exclusion criteria were intracranial stenting and intraarterial (i.a.) thrombolysis. Study groups were defined as patients developing vasospasms during EST (V+) and patients, who did not (V-). The study groups were compared in univariate analysis. Further, multivariable regression models were developed to predict the patients’ risk for developing vasospasms based on pre-identified potential prognostic factors. Secondary endpoint was clinical outcome as modified Rankin Scale (mRS) difference between pre-stroke mRS and discharge mRS (delta mRS).

**Results:** In total, 132 of 1768 patients (7.5 %) developed vasospasms during EST in this study cohort. Patients with vasospasms were younger and had a lower pre-stroke disability (as per mRS). Vasospasms were more likely to occur in ESTs with multiple thrombectomy attempts in total and after several stent retriever maneuvers. Factors with predictive value for developing vasospasms were younger (OR = 0.967, 95%-CI = 0.96 - 0.98) and had a lower pre-stroke mRS (OR = 0.759, 95%-CI = 0.63 - 0.91). The prediction model incorporating patient age, pre-stroke mRS, stent retriever thrombectomy attempts, and total attempts as prognostic factors was found to predict vasospasms with decent accuracy (AUC = 0.714, 95%-CI = 0.709-0.720). V+ patients showed higher delta mRS (median (Q1 - Q3); V-: 2 (1-3) vs. V+: 2(1-4); p = 0.014).

**Conclusion:** This study shows that vasospasms are a common complication in EST affecting younger and previously healthier patients and are more likely after multiple stent retriever thrombectomy attempts. As independent predictors patient age, pre-stroke mRS, thrombectomy maneuvers and stent-retriever attempts predict the occurrence of vasospasms during EST with decent accuracy.

## INTRODUCTION

The description of vasospasms as a complication in endovascular stroke treatment is as old as the therapy itself. Gupta et al. described arterial vasospasms after stent-retriever thrombectomy in 2009 [1]. Since 2015, with the establishment of endovascular stroke treatment (EST) as a first-line therapy for acute ischemic stroke, more research emerges concerning the management of procedure failure and interventional complications [2,3]. In the early randomized multicenter studies of 2015, only REVASCAT and SWIFT PRIME reported on vasospasms with fourteen cases of vasospasms in the intervention group of REVASCAT (14/103, 13.6 %), four of which required treatment by application of vasodilating medication, and four reported cases of vasospasm (4/98, 4 %) in the intervention group of SWIFT PRIME [4,5]. More studies showed the occurrence of intracranial vasospasm in 3% to 19% of EST cases [6,7]. A study by Akins et al. describes vasospasms as a very common complication during EST in 16% of all cases [8].

Vasospasms during EST are typically considered to be caused by the mechanical irritation of the vessel by thrombectomy attempts using stent retrievers, aspiration catheters or both. McTaggart et al. recommend desisting from using a stent retriever when a vasospasm of the target vessel is detected as they suppose that vasospasms might be induced and reinforced by using stent retrievers [9]. A single institution study analysis of 240 patients found a rate of vasospasms of 2.1% (1/47 patients) in EST without the use of stent retrievers and a rate of 11.3% (17/151 patients) with the use of a stent retriever [10]. In SWIFT PRIME the authors regard the occurrence of intraprocedural vasospasm as a nonserious and transient adverse event without further clinical sequelae in the four cases they observed in their study [11]. Emprechtinger et al. consider vasospasms during EST as a possible cause for recurrent stroke [4].

In patients with subarachnoid hemorrhage, it is well known that especially younger patients are prone to develop vasospasms [12]. It was found that patient age < 50 years is associated with a five-fold greater risk of vasospasm compared with older patients. The pathophysiological reasons for the phenomenon are unclear, but vascular risk factors in older patients, influencing the receptiveness of vessel walls to vasospasm triggers, are discussed as potentially important factors. In patients suffering from subarachnoid hemorrhage the occurrence of vasospasms has clearly been established as one of the major determinants for unfavorable outcome.

In patients receiving EST due to acute intracranial vessel occlusion, this age dependency or other predictors for the occurrence of vasospasms during EST are still undefined. The influence of vasospasms on the clinical outcome is also still not understood. The aim of this study was to determine independent predictors for vasospasms in acute ischemic stroke patients and anterior circulation EST as well as define the influence of vasospasms during EST on the clinical outcome.

## METHODS

This is a retrospective, single-center clinical study. All consecutive patients with acute ischemic stroke in the anterior circulation and at least one intracranial thrombectomy attempt were selected from an institutional review-board approved prospective database of a tertiary stroke center in Germany treated between January 2015 to December 2021. Ethics committee of Medical Faculty, University of Heidelberg (Germany) gave ethical approval for this work.

### Patient selection and study groups

Inclusion criteria for this study were an intracranial target vessel occlusion in the anterior circulation and at least one thrombectomy attempt. Exclusion criteria were i) intracranial stenting ii) intraarterial thrombolysis – see figure 1.

**Figure 1.**
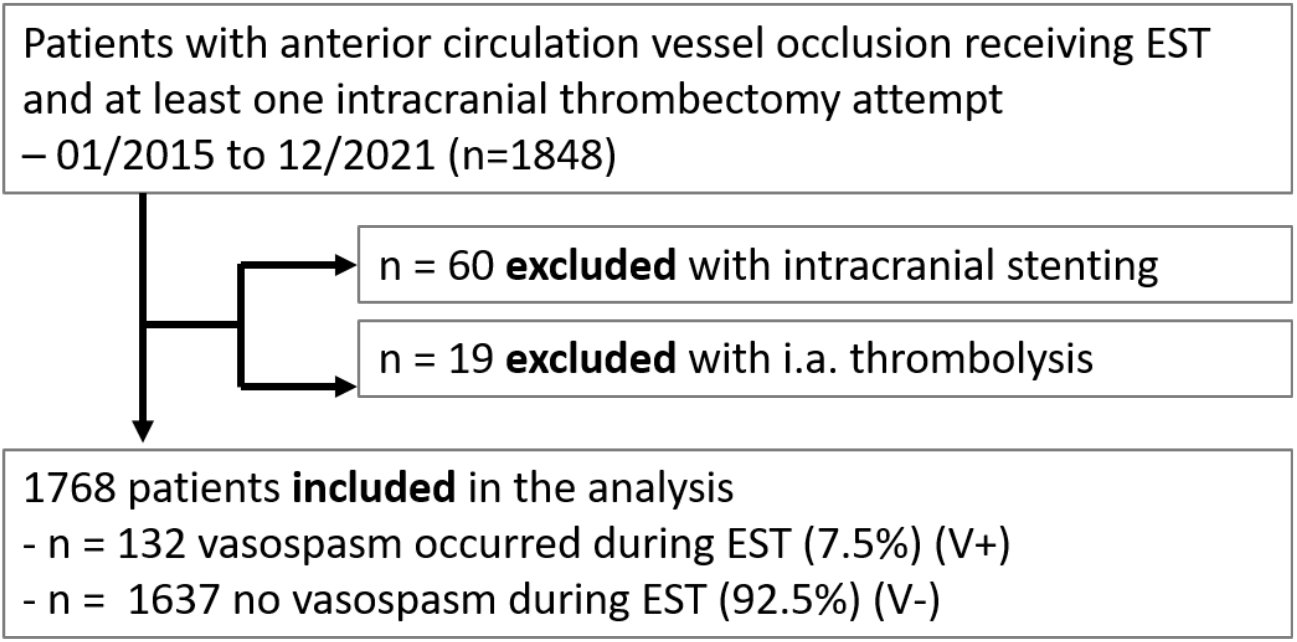
*Patient selection* for comparison of patients with vasospasm (V+) during endovascular stroke treatment vs. no vasospasm (V-) during endovascular stroke treatment; EST, endovascular stroke treatment.

The two study groups were defined according to vasospasms during EST (V+) and no vasospasms during EST (V-). The study groups were compared in univariate analysis comprising relevant clinical and imaging data.

Vasospasms were defined as a concentric vessel constriction (> 70 % stenosis) after EST maneuver [13]. At the study facility, the appearance of vasospasms prompts local infusion of intra-arterial nimodipine via microcatheter or via the intermediate catheter flush. Nimodipine is not routinely added to infusions during EST in the study facility.

### Statistical analysis

Patients’ demographics and clinical examination data were assessed descriptively in both study groups. Continuous variables were described using the median, Q1 and Q3. For categorical variables absolute and relative frequencies were provided.

In addition, a two-sided Welch’s two-sample t-test and a test of homogeneity (the Fisher’s exact test or Boschloo’s test) were performed for continuous and categorical variables, respectively, to assess whether both groups significantly differed regarding the variables of interest at the 5% level. P-values of the tests were reported alongside 95% confidence intervals for the mean difference (Welch’s two-sample t-test) and the proportion difference (Fisher’s exact test) or odds ratio (Boschloo’s test). All p-values were interpreted descriptively. A multivariate analysis was conducted to evaluate whether and to which extent pre-identified potential prognostic factors impacts the odds for developing vasospasms. Missing values in the data were imputed ten times using the predictive mean matching method. For each imputed data set, a variable selection was performed using the elastic net regression with repeated five-folds cross-validation [14]. The set of candidate prognostic factors were the patient age, pre-stroke mRS, stent retriever thrombectomy attempts, total thrombectomy attempts, coronary heart disease, hypertonia, number of aspirations maneuvers, patient sex, onset to puncture (in min), closure localization, and stenting. At each iteration of the cross-validation procedure, the model and per extension, the subset of prognostic factors which maximized the predictive performance as measured by the Area Under the Curve (AUC) were selected. For each potential prognostic factor, the relative frequency of its inclusion in the best subset of prognostic factors over all 500 cross-validation iterations (100 iterations of five-folds cross validation) and all ten imputed data sets was then calculated allowing to identify the prognostic factors with the highest predictive value.

Further, to evaluate how the selected prognostic factors impact the odds of appearance of vasospasms, the original data were imputed ten times again using predictive mean matching and a logistic regression model fitted to each imputed data set with the selected sets of prognostic factors as independent variables. The resulting regression coefficients of all ten logistic regression models were pooled together following the Rubin’s rules and interpreted [15].

The predictive performance of the fitted regression models was assessed separately using the AUC. Hereby, a logistic regression analysis with repeated five-folds cross-validation was conducted on each imputed data set. For each imputed data set, the obtained AUC values were averaged over all five cross-validation folds and then over 100 cross-validation iterations. The aggregated AUC values were then logit-transformed, pooled over all imputed data sets using the Rubin’s rules and back-transformed to the original scale to obtain a single AUC value and the corresponding 95% confidence interval for each set of selected predictors.

Secondary endpoint was the modified Rankin Scale (mRS) difference before the stroke mRS and at discharge (delta mRS) and was investigated by means of a two-sided Welch’s two-sample t-test. The significance level was set to p=0.05. A p-value was reported alongside a 95% CI for the mean delta mRS difference between both groups. Statistical analysis was conducted using R version 4.1.3.

### Data Acquisition

Source data were generated from a prospectively collected stroke database. Additionally, all data included in the present analysis were validated retrospectively to minimize incorrect or missing data (JJ, CW). The EST’s angiographic imaging was reviewed to assess the occurrence and potential resolution of vasospasms (JJ, CW). Clinical data were assessed by the treating neurologist during acute hospital stay.

## RESULTS

In this study cohort of 1768 patients with acute ischemic stroke, target vessel occlusion in the anterior circulation and EST, 132 patients (7.5%) developed vasospasms (V+) during EST. In 113 cases (6.3%) vasospasms were located intracranially, in 29 cases (1.6%) extracranially, and in 7 cases (0.4%) both intra- and extracranially.

Patients developing vasospasms were younger (age in years: median, IQR; V-: 78, 67 – 84 vs. V+: 66, 57 – 76; p < 0.001) and healthier with a lower pre-stroke mRS (median, IQR; V-: 1, 0 – 2 vs. V+: 0, 0 – 1; p < 0.001). Vasospasm patients had less comorbidities such as coronary artery disease (n, %; V-: 416, 25.4% vs. V+: 20, 15.2%, p = 0.011), arterial hypertension, atrial fibrillation and diabetes type II – see table 1.

**Table 1.**
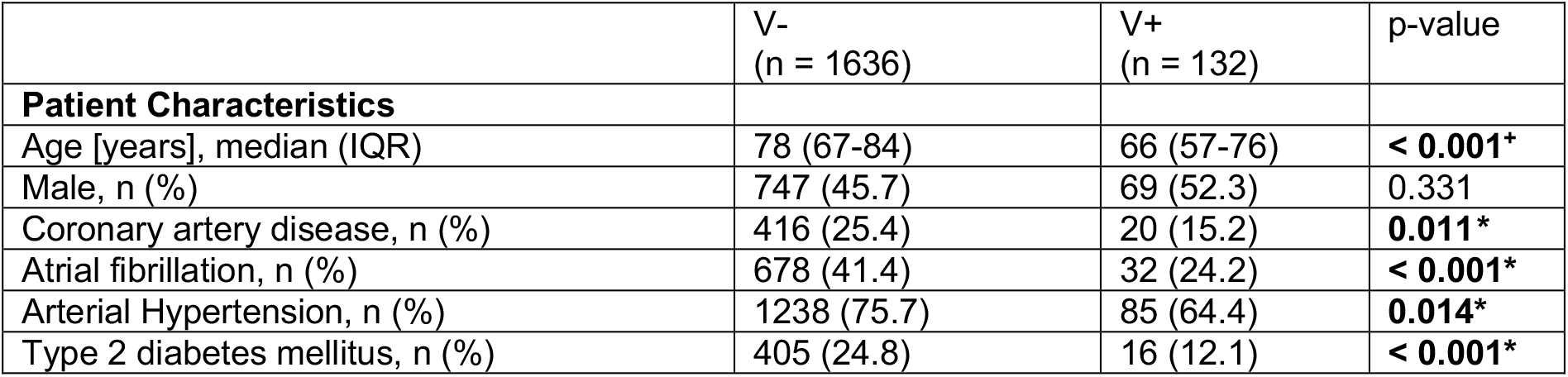
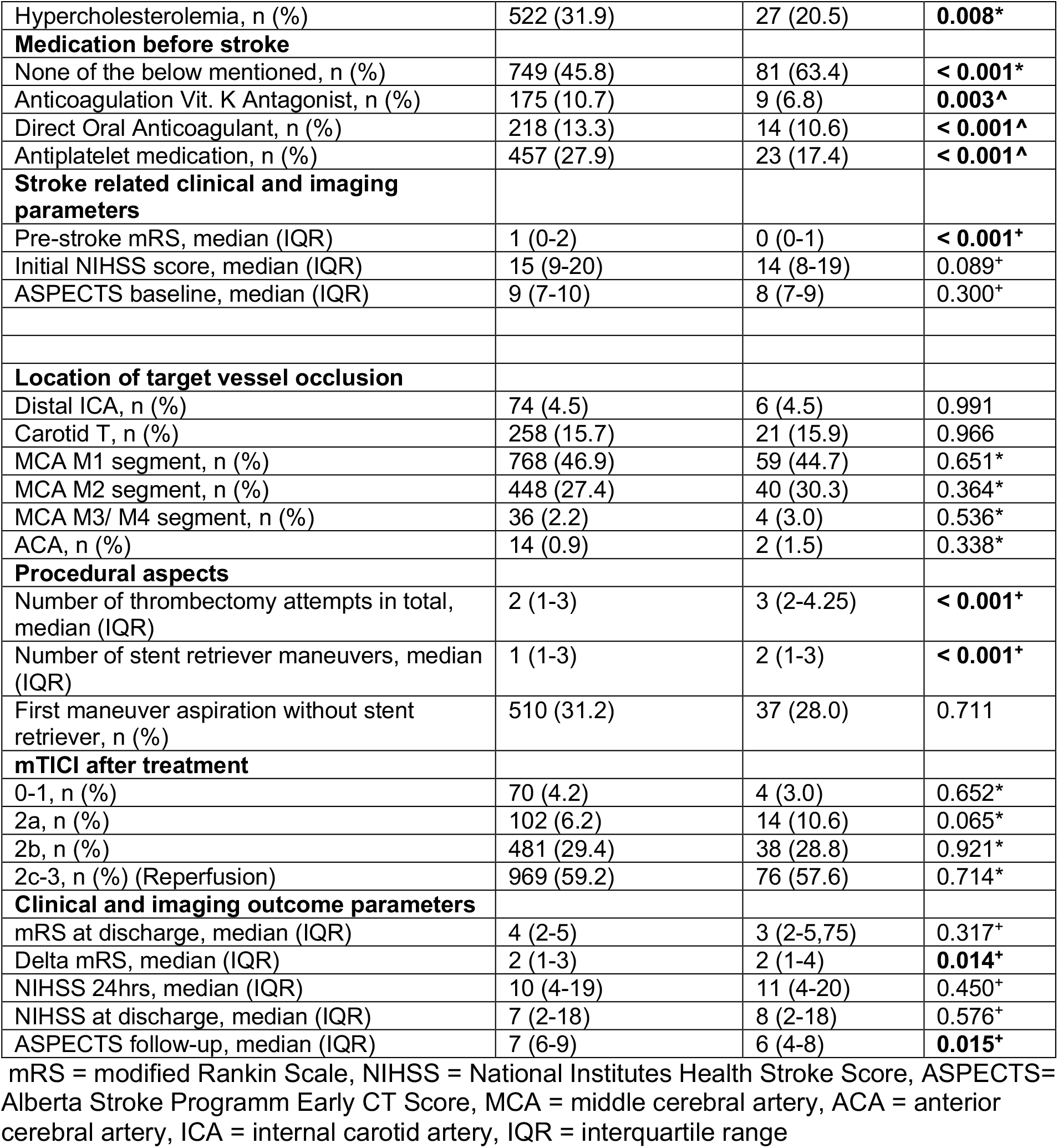
Group comparison of patients with vasospasm (V+) and no vasospasms (V-) during endovascular stroke treatment. Bold values are statistically significant p-values (< 0.05); Chi-Square-Test. ^+^ Mann-Whitney-U-Test. * Fisher exact test. ^Fisher-Boschloo test.

The target vessel occlusion site did not differ significantly between the two groups – see table 1. More thrombectomy attempts (contact aspiration or stent-retriever thrombectomy) were performed in V+ patients (median, IQR; V-: 2, 1-3 vs. V+: 3, 2 – 4.25; p < 0.001) as well as more stent-retriever maneuvers (median, IQR; V-: 1, 1-3 vs. V+: 2, 1 – 3; p < 0.001). The initial technical thrombectomy approach – namely contact aspiration or stent retriever thrombectomy – did not influence the occurrence of vasospasms (first maneuver contact aspiration V-: 31.2% vs. V+: 28.0%, p = 0.711). The prevalence of tandem occlusions and Stent-assisted PTA for extracranial high grade ICA stenosis or occlusion was comparable between study groups (n, %; V-: 943, 57.4% vs. V+: 67, 50.8%, p = 0.133). Vasospasms were treated by local standard operating procedure protocol with intra-arterial nimodipin (105 patients, 77.8% of patients with vasospasm) resulting in near-complete or complete resolution of vasospasms in the majority of the cases (91 patients of 105 patients, 86.7 %) and a comparable mTICI score at the end of the EST compared to patients without vasospasms (mTICI 2b-3, V-: 88.6% vs. V+: 86.4%, p = 0.824).

Further, elastic net regression analysis revealed that with a 100% selection probability over 5000 iterations, age, pre-stroke mRS, stent retriever maneuvers, and total number of thrombectomy attempts, have the highest predictive value for developing vasospasms. As shown in table 2, the logistic regression analysis found patient age and pre-stroke mRS to be significant prognostic factors for developing vasospasms during EST in the anterior circulation in the study cohort (age: OR = 0.967, 95%-CI = 0.96-0.98, pre-stroke mRS OR = 0.759, 95%-CI = 0.63-0.91). It showed that for two patients with one-year age difference, the odds of vasospasms during EST are 3.2% higher for the younger patient. Further calculation revealed that the risk to develop vasospasms during EST doubles for a patient who is 20 years younger compared to an average patient in this study cohort – see figure 2. Regarding pre-stroke mRS, it appears that for two patients with one-unit difference in pre-stroke mRS score, the odds of vasospasms are 24.1% higher for the patient with the lower score.

**Table 2.** Logistic regression analysis with age, pre-stroke mRS, total number of thrombectomy attempts and stent-retriever maneuvers as prognostic factors. Pooled regression coefficients over 10 imputations.

| Variable | Selection probability |
| --- | --- |
| **Patient age** | 1.000 |
| **Pre-stroke modified Rankin Scale Score** | 1.000 |
| **Number of total thrombectomy maneuvers** | 1.000 |
| **Stent-retriever thrombectomy maneuvers** | 1.000 |
| Coronary artery disease | 0.769 |
| Chronic hypertension | 0.105 |
| Contact aspiration thrombectomy maneuvers | 0.102 |
| Sex | 0.041 |
| Stroke symptom onset to groin puncture time in minutes | 0.022 |
| Intracranial Target Vessel Occlusion | 0.014 |
| Tandem occlusion with stent-assisted PTA of the extracranial carotid artery | 0.014 |

**Figure 2.**
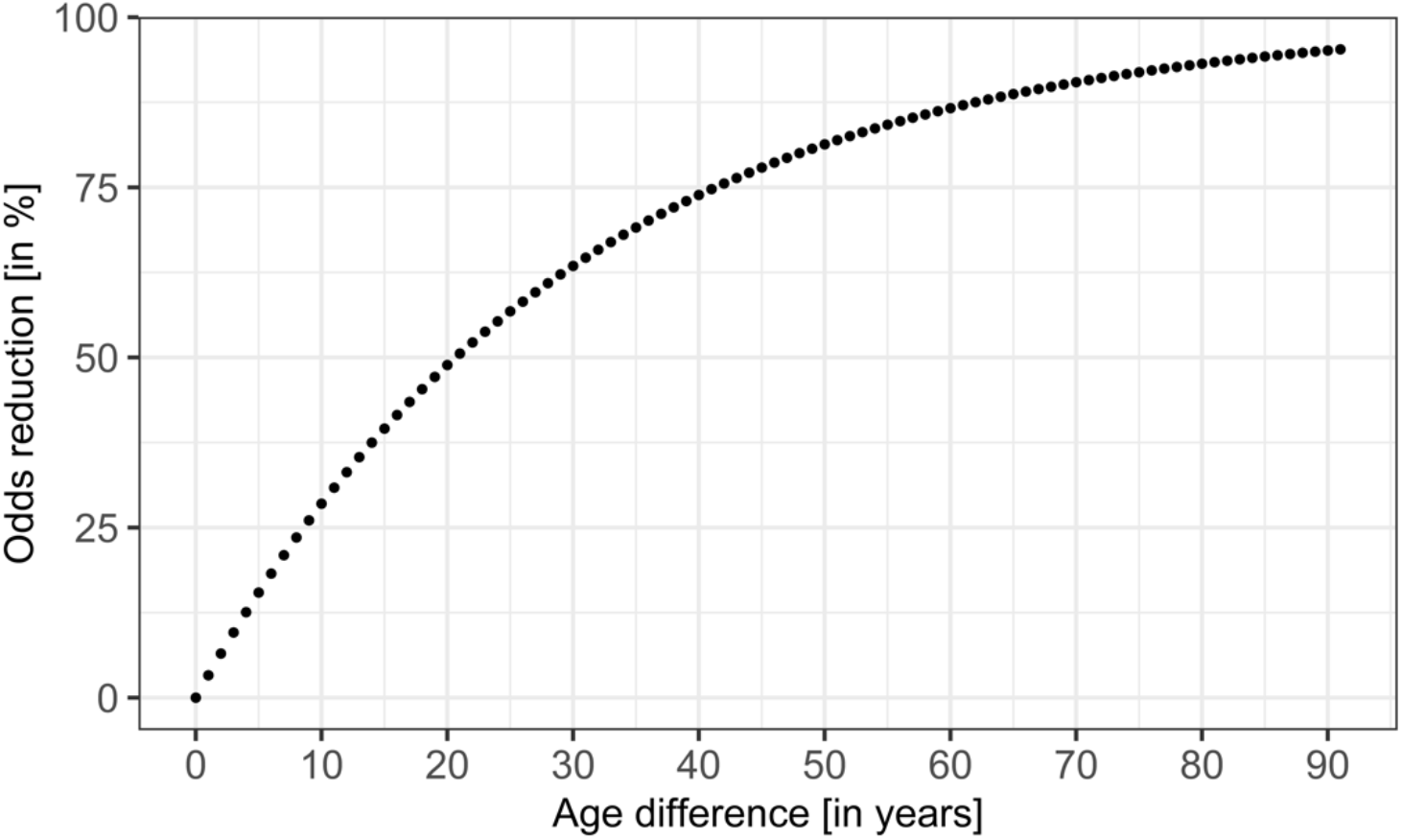
Relationship between patients’ age difference in years and odds reduction for developing vasospasms.

Although V+ patients show less comorbidities like coronary artery disease (CAD), including CAD in the model leads to only minimal change in the regression coefficients (see table 3).

**Table 3.** Logistic regression analysis with age, pre-stroke mRS, total number of thrombectomy attempts, stent-retriever maneuvers and coronary artery disease as prognostic factors. Pooled regression coefficients over 10 imputations.

|  | OR | SE | p-value | Lower CI | Upper CI |
| --- | --- | --- | --- | --- | --- |
| Intercept | 0.750 | 0.454 | 0.526 | 0.308 | 1.827 |
| Age | 0.967 | 0.007 | < 0.001 | 0.955 | 0.980 |
| Pre-stroke modified Rankin Scale | 0.759 | 0.094 | 0.003 | 0.631 | 0.912 |
| Stent retriever thrombectomy attempts | 1.092 | 0.087 | 0.311 | 0.921 | 1.296 |
| Total number of thrombectomy attempts | 1.066 | 0.071 | 0.374 | 0.926 | 1.226 |

Moreover, a prediction model comprising patient age, pre-stroke mRS, total number of thrombectomy attempts and stent-retriever maneuvers was found to predict the odds for developing vasospasms with a decent accuracy (AUC = 0.714, CI = 0.709-0.720). The calibration plot in figure 3 shows that most of the predicted probabilities are at the lower end and the model seems to be well calibrated in that area. However, high underpredictions and overpredictions can be observed at higher prediction probabilities. Although the overall calibration as measured by the calibration intercept is zero (intercept = 0, CI = -0.18-0.18), the calibration slope is lower than 1 (calibration slope = 0.93, CI = 0.69-1.17) suggesting that the estimated risks are too high for patients who are at high risk and too low for patients who are at low risk. Including CAD in the model did not improve its prediction accuracy (AUC = 0.713, CI = 0.708-0.718) – see also table 4. The resulting calibration plot was almost identical in shape, and the calibration intercept and slope only changed minimally (see figure 4).

**Figure 3.**
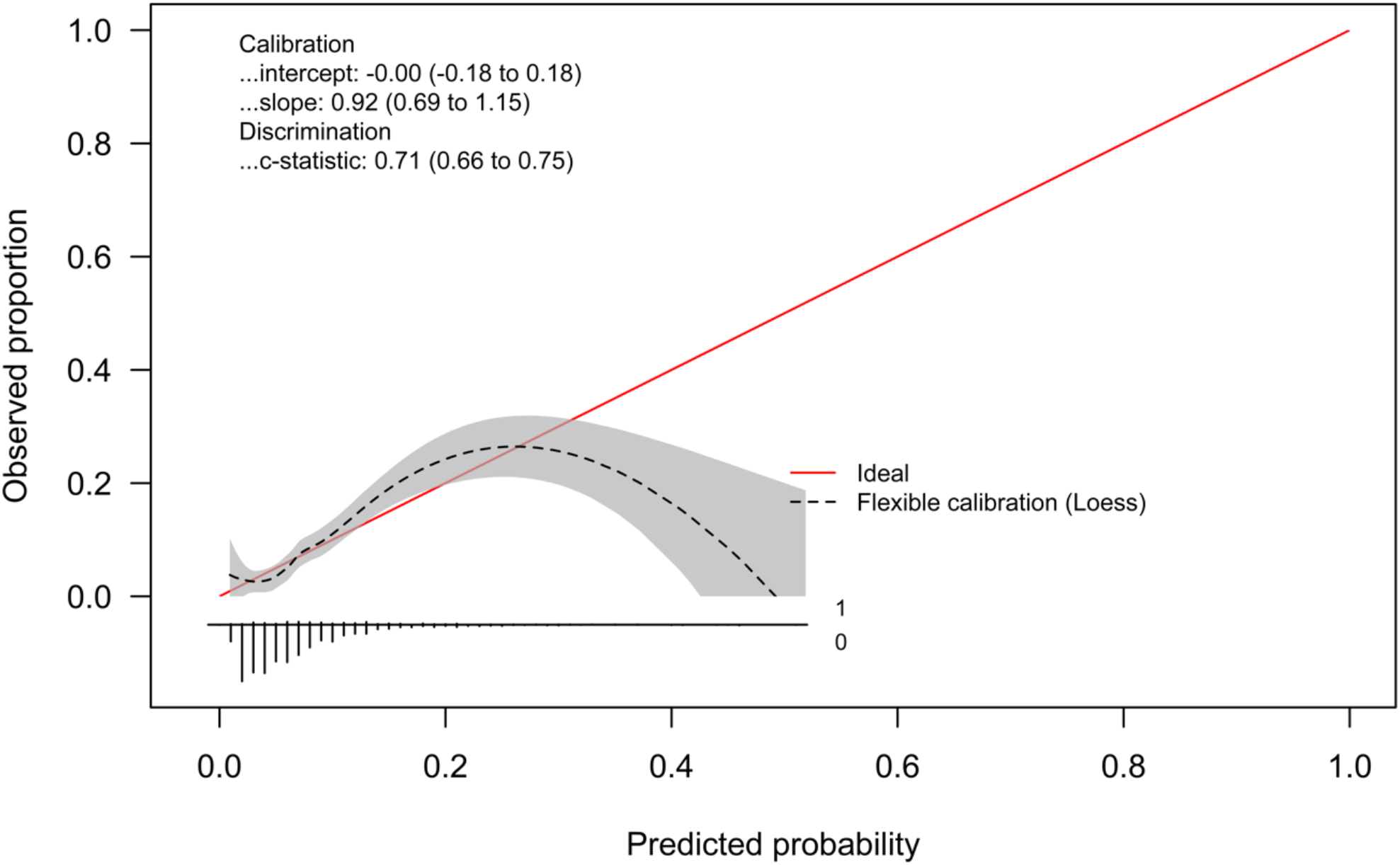
Logistic regression calibration plot for the prediction regression model with age, pre-stroke mRS, total number of thrombectomy attempts and stent-retriever maneuvers (event rate=7%).

**Figure 4.**
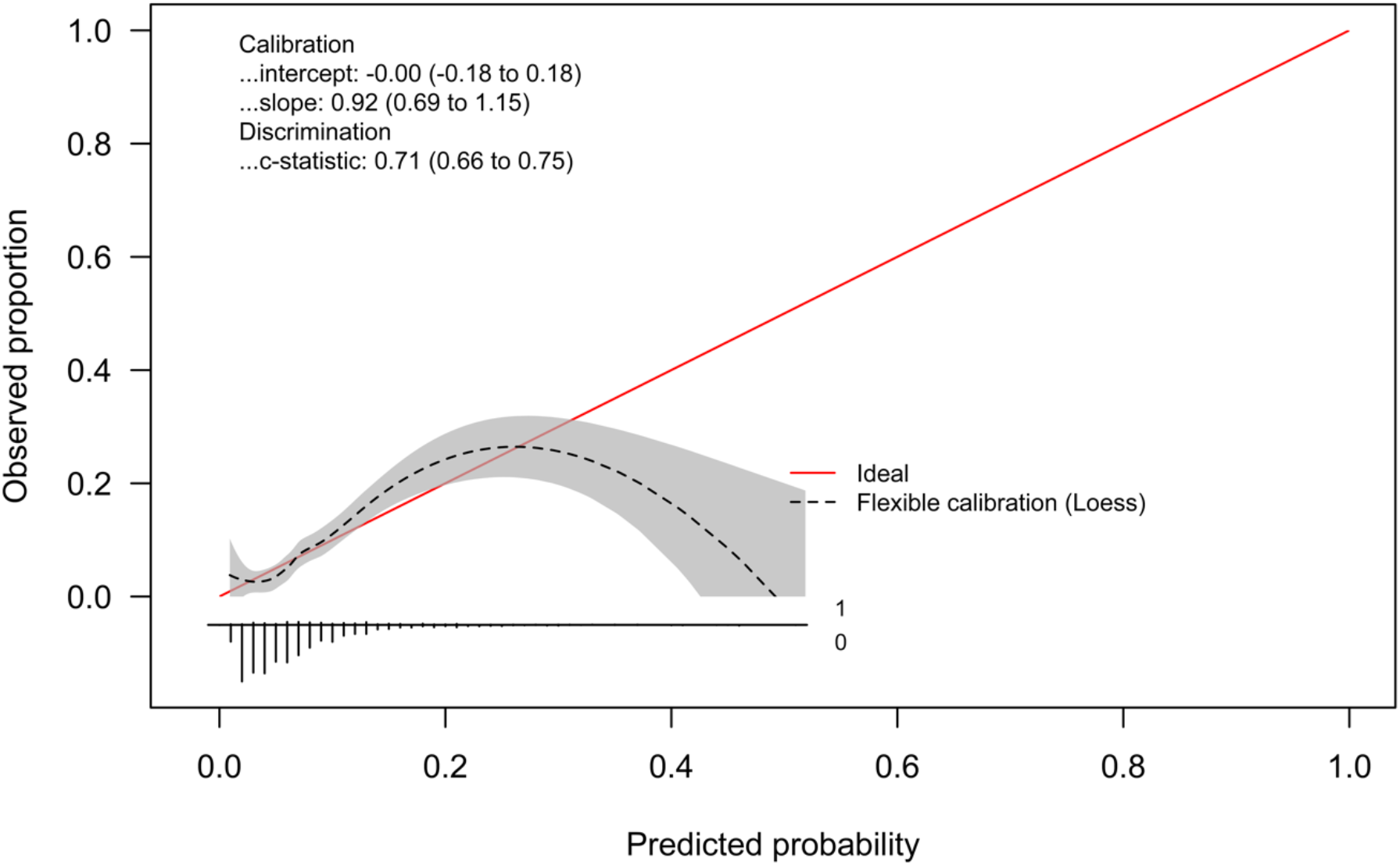
Logistic regression calibration plot for the prediction regression model with age, pre-stroke mRS, total number of thrombectomy attempts, stent-retriever maneuvers and coronary artery disease (event rate=7%).

**Table 4.** Prediction models for vasospasms during endovascular stroke treatment. mRS = modified Rankin Scale, SR = Stent Retriever, CAD = Coronary Artery Disease, AUC= Area Under the Curve, CI = Confidence Interval

| Predictor set | AUC | Lower CI | Upper CI |
| --- | --- | --- | --- |
| Age + pre-stroke mRS + SRmaneuver + total maneuver count | 0.714 | 0.709 | 0.720 |
| Age + pre-stroke mRS + SRmaneuver + total maneuver count + CAD | 0.713 | 0.708 | 0.718 |

As for the secondary study endpoint, the difference between discharge mRS and pre-stroke mRS was significantly higher for V+ patients (median (IQR): V-: 2 (1-3) vs. V+: 2 (1-4), p = 0.014).

## DISCUSSION

Vasospasms are a common complication during EST in the anterior circulation with 7.5% in this study cohort, which is in line with previous reports ranging from 3% to 23% [6,7]. As a main finding, this study shows that vasospasms rather occur in younger patients and are to be expected in ESTs with more thrombectomy maneuvers in total and especially after multiple stent retriever maneuvers. The first thrombectomy technique (contact aspiration or stent retriever thrombectomy under aspiration) did not differ between the two study groups of patients without vasospasms (V-) and patients with vasospasms (V+).

To our knowledge, this is the first study presenting predictors for vasospasms during EST, namely patient age, pre-stroke mRS, thrombectomy maneuvers in total and stent-retriever maneuvers. They can predict the risk of vasospasms during EST with decent accuracy (AUC = 0.718). According to this study, the incidence of vasospasm doubles with an age difference of 20 years for the younger patient.

The age dependency of vasospasms is well known for patients with subarachnoid hemorrhage [12,16] but has not been shown for stroke patients receiving EST yet. The younger patients in the V+ study group showed less comorbidities like coronary artery disease, which we supposed might be correlated to intracranial arteriosclerosis and decreased vessel wall elasticity and the propensity for the development of vasospasm. However, coronary artery disease according to our prediction analysis did not play a role in the development of vasospasms during EST.

Furthermore, with every stent retriever maneuver the risk of vasospasm increases by 6%. Previous studies reported a higher rate of vasospasm in patients treated by stent retriever thrombectomy compared to aspiration thrombectomy or treated with more stent retriever maneuvers [9,10]. As for the management of complications during EST, our study facility follows the recommendation of using calcium channel blockers, such as nimodipine. Nimodipine is a dihydropyridine agent that blocks voltage-gated calcium channels and has a dilatory effect on arterial smooth muscle with a half-life of about nine hours [17]. Pilgram-Pastor et al. 2021 suggest a rate of 0.5 - 1 mg nimodipine i.a. over several minutes and points out the risk of hypotension and steal phenomena [3]. Others suggest the use of 2 – 5 mg nicardipine i.a. as soon as a tendency to focal vasospasm can be detected [10]. In our study nimodipine efficiently resolves vasospasms caused by EST in 86.7% of cases, but its influence on infarct growth and stroke outcome is still unknown.

Regarding outcome measures, this study shows lower ASPECTS scores, corresponding to larger ischemic infarcts, after EST for patients with vasospasms. This seems surprising as patients with vasospasms were treated immediately with nimodipine leading to a restored vessel diameter in the majority of cases and leading to comparable reperfusion rates in the V+ and V-study groups. Supposedly this is related to vessel re-occlusion after EST in at least some of the vasospasm patients. As we do not routinely perform imaging angiographies after EST in our institution, we cannot verify this hypothesis and think it should be investigated in further studies. Also, Emprechtinger et al. discussed that recurrent stroke could be caused by vasospasms induced by EST [4]. Another hypothesis is that vasospasm comprises macro- and microcirculation, whereas treatment with intra-arterial drug delivery has an effect merely on larger vessels and not on the microcirculation. Thus, there is still impaired blood flow with subsequent infarction of tissue.

Compatible with our imaging finding of larger infarcts in patients with vasospasms, these patients also showed a worse outcome regarding their mRS at discharge in relation to their initially lower mRS before stroke onset (delta mRS) compared to patients without vasospasms. Consequently, the increased infarct volumes and worse short-term outcome should prompt more studies on the monitoring and management of patients with vasospasms after the initial EST.

Limitations of this study are related to the single-center, retrospective character. Especially the differing operating procedures of stroke centers might result in different occurrence rates of vasospasms and related outcome.

## CONCLUSION

Vasospasms during EST in the anterior circulation represent a severe complication affecting mainly the younger and previously healthier stroke patient resulting in larger infarcts and worse short-term outcome. Predictors for vasospasms are patient age, pre-stroke mRS, number of total thrombectomy maneuvers and stent-retriever maneuvers, that predict the occurrence of vasospasms during EST with decent accuracy.

## Data Availability

All data produced in the present study are available upon reasonable request to the authors

## Appendix

